# How and when to end the COVID-19 lockdown: an optimisation approach

**DOI:** 10.1101/2020.04.29.20084517

**Authors:** Thomas Rawson, Tom Brewer, Dessislava Veltcheva, Chris Huntingford, Michael B. Bonsall

## Abstract

Countries around the world are in a state of lockdown to help limit the spread of SARS-CoV-2. However, as the number of new daily confirmed cases begins to decrease, governments must decide how to release their populations from quarantine as efficiently as possible without overwhelming their health services. We applied an optimal control framework to an adapted Susceptible-Exposure-Infection-Recovery (SEIR) model framework to investigate the efficacy of two potential lockdown release strategies, focusing on the UK population as a test case. To limit recurrent spread, we find that ending quarantine for the entire population simultaneously is a high-risk strategy, and that a gradual re-integration approach would be more reliable. Furthermore, to increase the number of people that can be first released, lockdown should not be ended until the number of new daily confirmed cases reaches a sufficiently low threshold.

We model a gradual release strategy by allowing different fractions of those in lockdown to re-enter the working non-quarantined population. Mathematical optimisation methods, combined with our adapted SEIR model, determine how to maximise those working while preventing the health service from being overwhelmed. **The optimal strategy is broadly found to be to release approximately half the population two-to-four weeks from the end of an initial infection peak, then wait another three-to-four months to allow for a second peak before releasing everyone else**. We also modelled an “on-off” strategy, of releasing everyone, but re-establishing lockdown if infections become too high. We conclude that the worst-case scenario of a gradual release is more manageable than the worst-case scenario of an on-off strategy, and caution against lockdown-release strategies based on a threshold-dependent on-off mechanism.

The two quantities most critical in determining the optimal solution are transmission rate and the recovery rate, where the latter is defined as the fraction of infected people in any given day that then become classed as recovered. We suggest that the accurate identification of these values is of particular importance to the ongoing monitoring of the pandemic.

## Introduction

### History of SARS-CoV-2 to date

Severe acute respiratory syndrome coronavirus 2 (SARS-CoV-2) is a novel coronavirus that has provoked the global pandemic of COVID-19. First reported in the city of Wuhan, China, its emergence quickly triggered a ‘lockdown’ within Wuhan and the surrounding cities^1^, requiring people to remain at home, only leaving for essential journeys. Since then the virus has spread rapidly worldwide, leading the World Health Organization to declare a global pandemic on the 11th of March. Globally, the outbreak has spread to 210 countries and territories, with 3,007,194 confirmed cases, 207,265 deaths and 883,298 recovered individuals, as of the 27th of April. (Viewed on April 27, 2020, 10:42 GMT^2,3^).

Containment of the virus has proven challenging. Although some patients will require intensive care, others have unreported mild symptoms, with as many as 17.9% of infected individuals possibly being asymptomatic^4^. Those with compromised immunities, underlying health conditions, and of old age, are most at risk of developing acute respiratory distress syndrome (ARDS) and subsequent respiratory failure, necessitating the use of mechanical ventilators in a dedicated intensive care unit (ICU)^5^. This mass spread of infection, and increasing pressure on hospital capacity has led the UK to follow the example of neighbouring European countries by officially implementing a lockdown as of March 23rd. While this appears to have slowed the spread of infection, the cost to the economy of such measures is considerable, with the first 1.5 months of lockdown estimated to have cost the UK 3.4% of its GDP^6^. With a viable vaccine still several months or years away, lockdown measures will eventually need to be lifted, but this must be done without risk of overwhelming the health service. If all restrictions are lifted universally, this could trigger a rapid resurgence of infections and cause further death.

Here we consider a two-way balance which aims to (i) maximize the number of people able to work outside of lockdown, while (ii) ensuring that the number of people with COVID-19 requiring medical help at no point crosses a threshold beyond which hospitals are unable to cope. As an immediate termination of lockdown for all is likely to trigger a surge in infections, a graded easing of lockdown restrictions is likely required. The focus of our analysis is to understand the optimal pathway by which to release people as safely as possible back into to a general and growing non-quarantined set of workers.

### Mathematical Modelling

To understand how to restart the economy yet avoid the saturation of health services, we present decision-making as a problem in optimal control. To determine an optimal solution requires two definitions. The first is a system of process-based differential equations whose boundary conditions or other attributes can be varied by policy decisions. The second definition is an objective function metric, which depends on the balance and extent to which our two conditions are fulfilled. The aim is to solve the differential equations, and find decisions affecting their boundary conditions that are optimal and maximise the objective function. Our equation set is based on a standard Susceptible, Exposed, Infected, Recovered (SEIR) model framework^7^. Each of the four components has a modelled population, and as time evolves, people move through each class towards recovery (or death). The novel part of our analysis is that the SEIR equations are solved for two groups (i.e. communities). The first community is a non-quarantined group, and during the full lockdown, this represents the essential workers required to maintain health provision, or essential services. The second community is those in quarantine. The main distinction between the non-quarantine and quarantine groups is that, in the latter, lockdown causes a much lower rate of virus transmission.

SEIR-based equations are solved for non-quarantine and quarantine groups, connected by modelled release strategies from lockdown. That is, we allow different fractions of the quarantined group to move into the non-quarantined group, and at different times. For each potential strategy of movement between the two groups, an objective function is calculated - some metric describing the desirability of such a strategy. This is high when many people are removed from quarantine, as they are available to work - a desirable outcome. However, its value switches to a near-infinite negative should the health service threshold be crossed due to high infection numbers. Our model calculates the highest possible objective function (the optimal strategy) and returns the number of release dates, their time of occurrence, and the number of people at each time. For comparison, we perform parallel simulations, where we release all in quarantine to the non-quarantine pool, but allow the return to quarantine later if necessary, should infections risk exceeding the capacity of the health services. We herein refer to this as a lockdown “on-off release” strategy, and again find optimal timings and number of releases.

No mathematical model, especially for something as complicated as virus transmission and human behaviours, can make predictions accurate to within a small margin of error. However, models are especially useful in two circumstances, and that we exploit. First, although simulations may lack absolute precision, predictions will have some level of robustness. Such predictions give strong indications of expected responses to a range of different boundary conditions, i.e. alternative release scenarios. Numerical model flexibility and speed of operation enables “what if?” questions to be asked of alternative forms of graded lockdown release. Second, by repeated operation of a model, it is possible to scan across ranges of parameter values. After governments start to release people, changes to infection levels can be compared against ensembles of simulations with perturbed parameters. Data-model comparison allows selection of the most appropriate parameter value; an approach sometimes referred to as “adaptive learning”. The trajectory for that value becomes a more reliable forecast for the days and weeks beyond the available data. Evidence this approach is feasible is illustrated in data of infections in countries before and during a lockdown. Although there is substantial geographic variation, all curves have similar forms, amenable to parameterisation. Indeed, politicians have frequently described a mathematical functional form, with the expression “flattening the curve”, used to explain why lockdown is essential to avoid overwhelming health services.

Our aim is to generate dynamical predictions and help inform the debate as to future lockdown release options. Each simulation can be readily understood in terms of policy decisions, and mathematically this implies careful parameterisation.

Our model is parameter sparse, yet sufficiently complex to capture a broad range of options. Critically, each parameter is elated to understandable quantities characterising infection levels or lockdown decisions. A central part of our analysis, in the bsence of much knowledge of the SARS-CoV-2 virus, is to vary our fundamental parameters to determine their effect on the optimal strategy. This identification of sensitivity aids understanding and can identify research priorities crucial to enhancing our understanding and ability to manage the COVID-19 pandemic.

## Methods

### Model Framework

Our model considers two parallel SEIR (Susceptible, Exposed, Infected, Recovered) systems, one describing the spread of isease in the quarantined ‘lockdown’ population, and the other capturing the spread amongst those at work. Under full lockdown, the latter pool contains only front-line workers who are unable to adopt social-distancing measures. This is captured by the following system of ordinary differential equations:

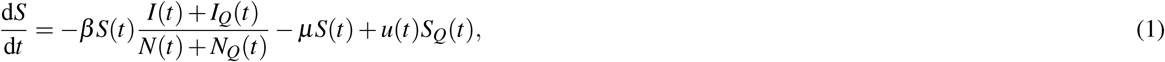

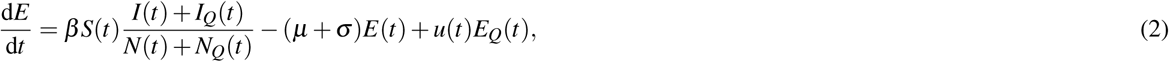

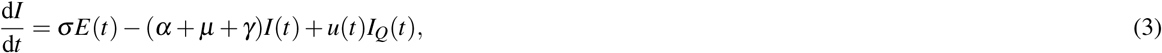

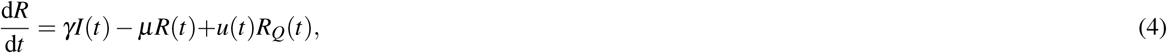

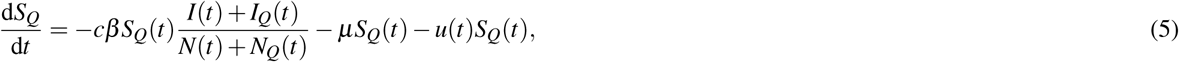

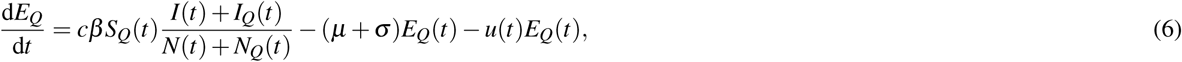

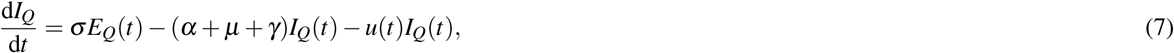

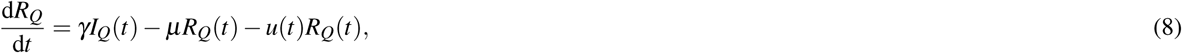

where *N*(*t*) = *S*(*t*) + *E*(*t*) + *I*(*t*) + *R*(*t*), and *N_Q_*(*t*) = *S_Q_*(*t*) + *E_Q_*(*t*) + *I_Q_*(*t*) + *R_Q_*(*t*), and the subscript Q denotes whether an individual is currently under lockdown conditions.

Our equations describe the movement of individuals through four stages, from being initially susceptible to the disease (*S*/*S_Q_*), contracting the disease but not yet being infectious (*E*/*E_Q_*), becoming infectious (*I*/*I_Q_*), and finally recovering from the disease (*R*/*R_Q_*), at which point we assume an individual becomes immune to future infections (Fig. 1). The function *u*(*t*) describes the release strategy employed, controlling the movement of individuals between the ‘quarantined’ and ‘released’ groups.

**Figure 1.**
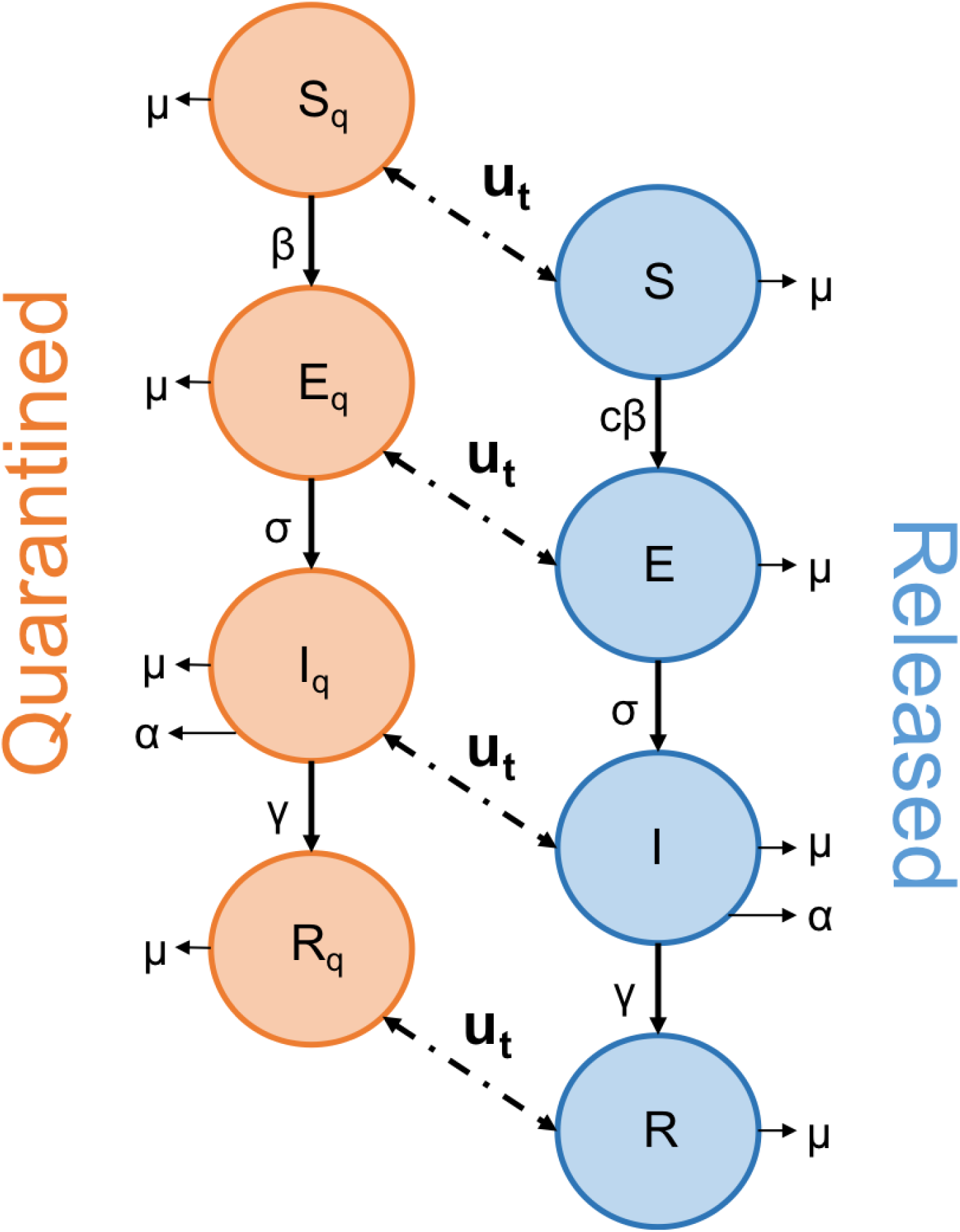
Schematic diagram depicting the movement of individuals through the SEIR network. The function u describes the action of the strategy employed to end lockdown, as people are released from the quarantined group. The arrows linking the two groups operate in both directions, to allow for any “on-off” strategy where people are returned to quarantine.

The lowercase Greek letters in equations (1) - (8) represent our rate parameters. Firstly, *β* represents the transmission rate of the disease. Significant work early in the pandemic used available data to quantify the rate of SARS-CoV-2 transmission and a range of estimates have already been reported in the literature. Kucharski et al. (2020)^8^ calculated an *R*_0_ of 1.15–4.77 when fitting to data from the initial outbreak in Wuhan. This corresponds in our case to a *β* ranging from roughly 0.25 - 1.06. Similarly, when fitting to data from the initial outbreak in Italy, Giordano et al. (2020)^9^ estimated a total transmission rate of 1.048, split between the four different infected classes they considered. These data-fit estimations risk failing to capture the impact of asymptomatic or unrecorded individuals, especially for the wider-ranging classes of our model, with a vision towards informing policy. For this reason our sensitivity analyses (below) also consider transmission rates up to twice as high as these values. Note that we consider the population of both *I* and *I_Q_* to impact the spread of disease, as the quarantined group are still assumed to occasionally mix with the population (for instance, when leaving their homes to shop for essential items). The parameter *c* is a scalar between 0 and 1 that captures how effective the self-isolation (i.e. lockdown) measures enforced are in reducing the the rate of SARS-CoV-2 transmission.

*μ* represents the natural, background death rate of the population regardless of the impact of COVID-19, and can have important implications for the strength of herd-immunity effects on disease dynamics, as this is the only mechanism in our model through which the recovered population is reduced. The parameter *α* represents the rate of death directly attributed to SARS-CoV-2. While the mortality rate of SARS-CoV-2 has been demonstrated to vary substantially between age classes^10–12^, in its current form our model does not incorporate age-structure and we therefore adopt an age-invariant mortality rate.

The parameter *σ* represents the incubation rate. The exposed population classes, *E*/*E_Q_*, capture the effect of the lag between people becoming infected (and incubating the disease for several days) and becoming infectious. Understanding the size of this effect is of great importance when assessing strategies in which a second lockdown may be enforced because efforts to monitor the subsequent spread of infection must consider the upcoming, but lagged, threat posed by the exposed class. Lastly, *γ* represents the recovery rate and describes how long individuals remain infectious.

In the present work, we used the population of the United Kingdom as an example to inform our initial proportion of the population in quarantine. Using Labour Force Survey data from 2018/19, the Institute for Fiscal Studies estimate that 7.1 million adults across the UK are in the set of key-worker guidelines set out by the UK government^13^. We define this 10.42% of the population as **not** currently being in lockdown, and initiate the model with the remaining population in lockdown. Initial numbers of individuals in each class were calculated using estimates presented by Flaxman et al. (2020)^14^, with the assumed values valid as of the 28th of March 2020.

All model variables, parameters and the values used for these are presented in Table 1. The full set of parameter estimates obtained, with links to the original sources, have been collated and made available in Appendix 1 to aid the modelling efforts of other research groups.

**Table 1.**
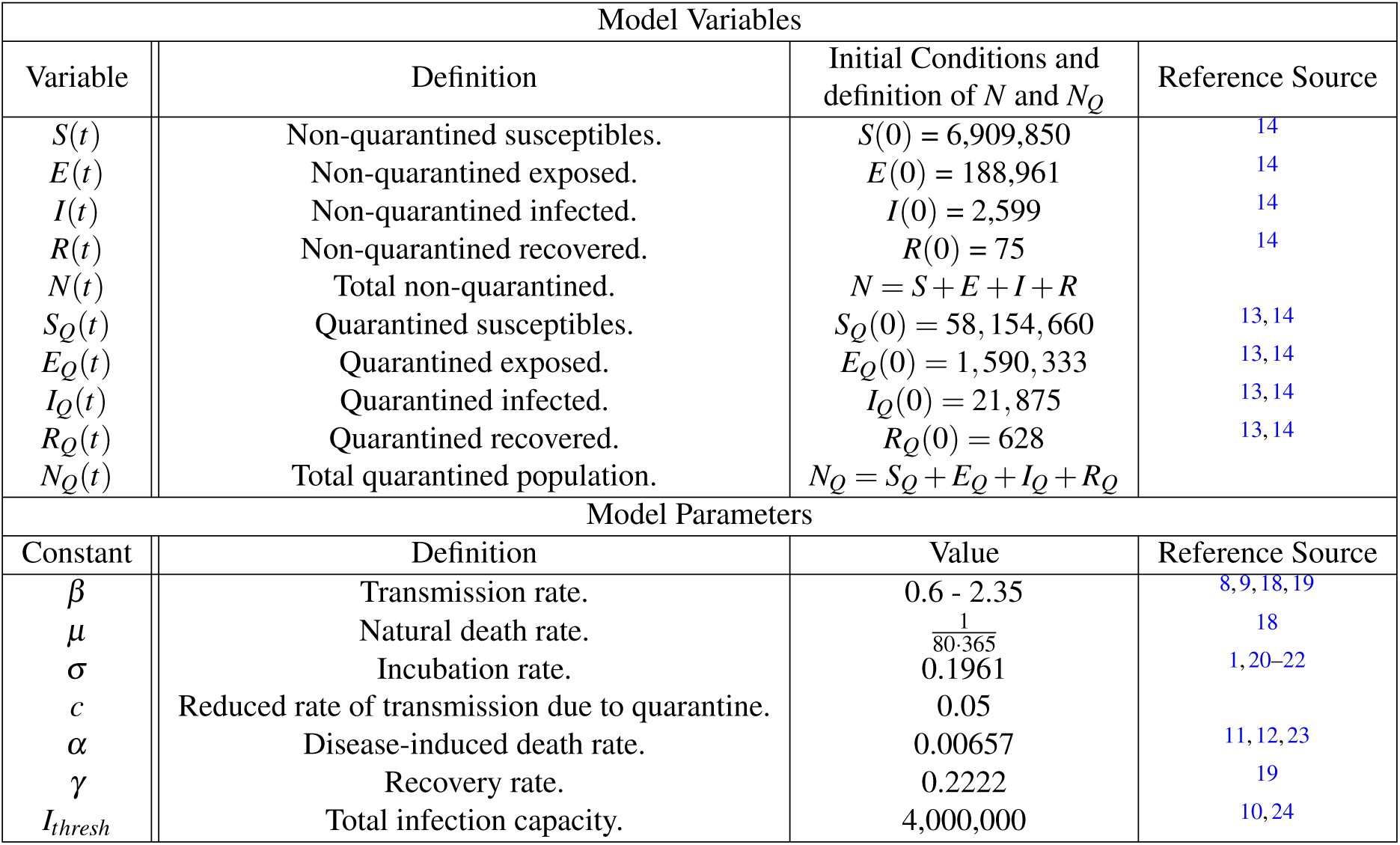
Definitions of the variables and parameters used in the SEIR model, of Equations (1) to (4) for the non-quarantined population, and Equations (5) to (8) for quarantined groups.

### Model Assumptions

Firstly, the extent and longevity of immunity to SARS-CoV-2, and its effect on the dynamics of the pandemic, remain open, high priority research questions^15,16^. Recent modelling efforts have, however, thus far shown little difference when incorporating the impact of waning immunity^9^. We therefore assume that, once developed, immunity provides complete and indefinite protection against SARS-CoV-2.

The parsimonious nature of our model was chosen to enhance the ease of interpretation of our results and, most importantly, to enable the model to be quickly adapted to non-UK populations. Different countries currently provide varying levels of epidemiological detail in their reporting of COVID-19 cases. By reducing the number of classes and parameters considered, our model is amenable to a wider range of countries and scenarios than the more specific model structures currently published^9,17^. The result of this modelling choice is that our system captures the broad-scale dynamics of the disease resulting from different lockdown exit-strategies rather than making accurate predictions of the number of infected individuals, which will require continuous, data-driven adaptations applied to our framework.

### Optimal Control

The primary challenge facing policy makers currently is in devising how to return the population to work most safely, ending the lockdown and its detrimental consequences on the economy. The objective is to release as many people from lockdown, as soon as possible, without overwhelming the health system with a subsequent resurgence of infections. This objective neatly fits the general framework of optimal control problems, a branch of mathematical study that seeks to maximise a certain objective functional through the use of available controls, while limited by constraints. In our model, the controls are the methods by which we release people from the quarantined class, described by the function *u*(*t*), and our constraint is our infection capacity, the maximum number of people our health system can effectively support at a given time. A solution is optimal if it returns the maximum number of people to work without breaking this constraint.

We consider two distinct strategies for ending the lockdown; a ‘gradual release’ strategy, whereby individuals are slowly, but permanently, released from quarantine in staggered waves until the entire population has been transferred from the quarantined class, and an ‘on-off release’ strategy, whereby the lockdown is lifted for the entire population simultaneously, but can subsequently be reinstated when necessary (the mathematical formulation of these strategies is outlined below). In each case, we seek to ensure that any strategy employed does not cause the total number of infected individuals (*I* + *I_Q_*) to surpass a certain threshold at any time. This threshold, *I_thresh_*, represents the maximum carrying capacity of the health service that cannot be exceeded. Ferguson et al. (2020) take the surge capacity of ICU beds in the UK to be 14 per 100,000 people^10^, equating to a total of 9240 ICU beds. They further note that as many as 30% of hospitalisations may require critical care. Combined with their estimate that 4.4% of all infections will require hospitalisations, this provides a range of values for *I_thresh_* to be considered in our sensitivity analysis, centred around an approximate threshold of 4,000,000 infected individuals.

Many formal optimal control approaches employ the use of “adjoint equations” to minimise the Hamiltonian of the ODE system. While we also pursued this approach, it requires a continuous-time form for the control function *u*(*t*), which (i) displayed extreme sensitivity in relation to any chosen objective functional, and (ii) is unlikely to be representative of lockdown release which, even if gradual, will still be managed with distinct groups of people leaving at different times. Our primary results presented in the following section are instead derived from an iterative process in which multiple different release times and portions of the population are trialled across various ranges, with the optimal choice being that which maximises our objective function. All code used to perform these optimal control approaches was performed in Matlab, and is available at: https://osf.io/hrt2k/.

### Gradual Release

A gradual release strategy aims to end the lockdown of the the public from quarantine through multiple staggered releases. Expressed mathematically, we seek to release *M*_1_ people at time *T*_1_, while ensuring that *I* + *I_Q_* < *I_thresh_* at all times. We iterate across a large mesh of potential values for *M*_1_ and *T*_1_, and for each trial we calculate the objective functional *C*_1_ = *M*_1_ – *T*_1_ – *J*(*I*, *I_Q_*), where *J*(*I*, *I_Q_*) is a penalty function that heavily penalises any iteration that results in *I*(*t*) +*I_Q_*(*t*) > *I_thresh_* for any *t*. Formally

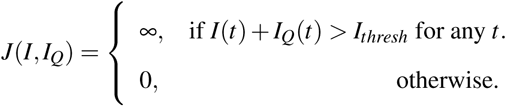

Therefore, the optimum choice of *M*_1_ and *T*_1_ are those which maximise *C*_1_. In short, this approach calculates how to release as many individuals as possible, as early as possible, without breaking the infection carrying capacity. After this optimum solution is found, a second release of *M*_2_ people at time *T*_2_ can be similarly calculated after the first release, if people still remain in quarantine.

To calculate these outputs, we used ode45, a fourth-order Runge-Kutta solver in Matlab, to solve the system of equations (1)-(8) using the initial conditions in Table 1 for *t* from 0 to *T*_1_. At this point we subtracted *M*_1_ individuals proportionally from *S_Q_, E_Q_, I_Q_* and *R_Q_* and added these to *S, E*, *I* and *R*. The system was then solved again from these new points for t from *T*_1_ to 400 days. To allow understanding of the effect of different values of some of the parameters presented in Table 1, we operate our model for a range of parameter values. Specifically, this was performed for a range of different transmission rates, *β*, infection thresholds, *I_thresh_*, and transmission reduction, *c*. Figure 2 depicts an illustrative example of a gradual release scenario.

**Figure 2.**
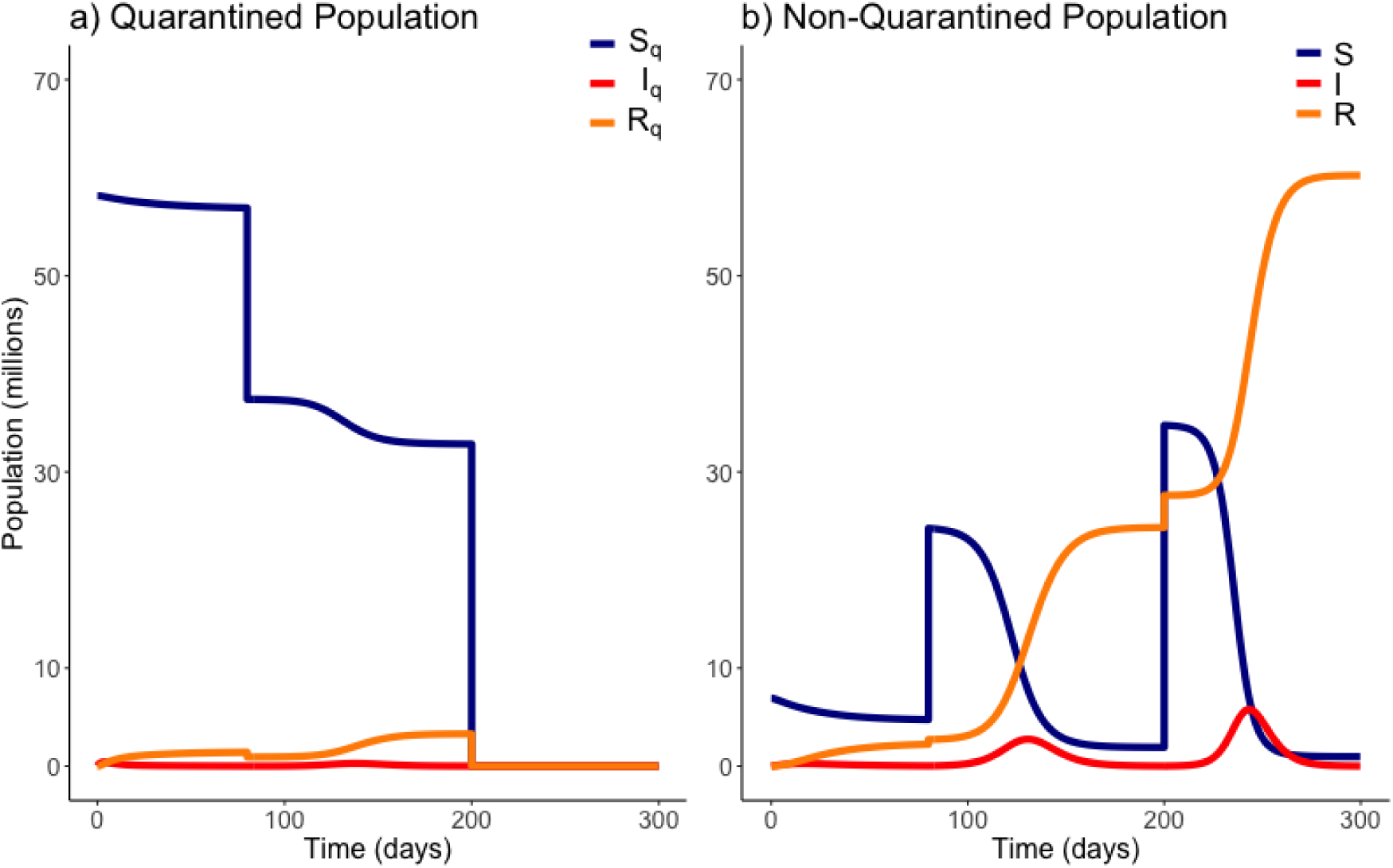
Example of a gradual release from quarantine. Here 20 million people are moved out of quarantine at *t =* 80 days, followed by the remaining population at *t* = 200 days. Variables *E_Q_* and *E* are not plotted. Model parameters are those of Table 1.

### On-Off Release

The “on-off” release strategy considers releasing the quarantined population all at once, with the aim to then return everyone to lockdown when required, should the number of infected exceed a threshold which threatens to overwhelm medical services. Formally, we seek *i* pairs of 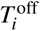 and 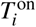, a time at which to end quarantine, and a time to re-instate it respectively. Consistent with the gradual release strategy, we iteratively trial multiple potential values of 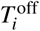 and 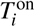 (where 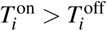).

For each choice of 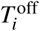 and 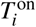 we calculate an objective functional 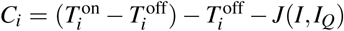where *J*(*I*, *I_Q_)* is as defined above. The optimum choice of 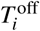 and 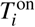 is the pair that maximises *C*. In short, we seek the longest possible duration out of quarantine, as soon as possible, without breaking the infection carrying capacity. We plot an example of an on-off release scenario in Figure 3 below.

**Figure 3.**
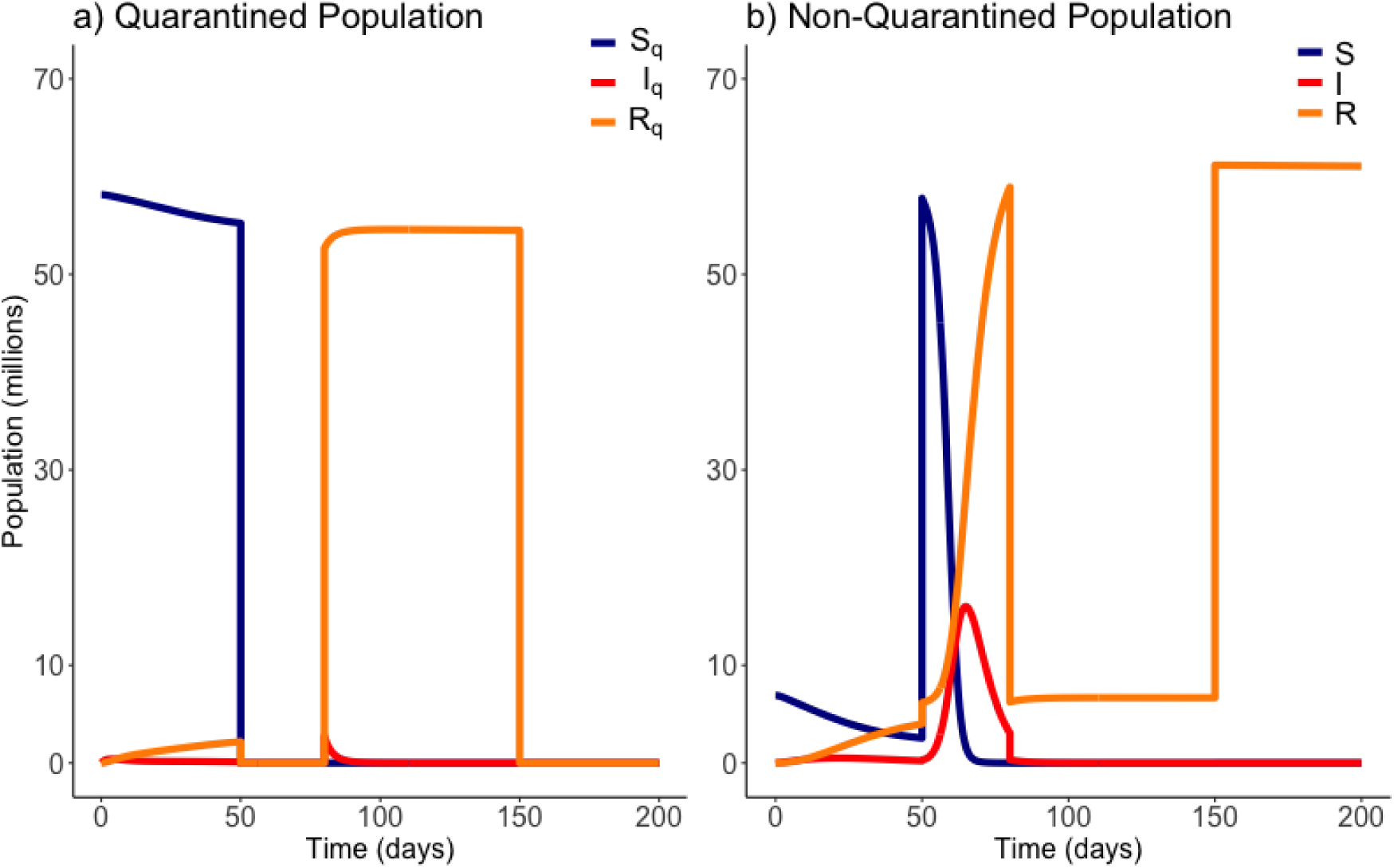
Example of an ‘on-off’ release from quarantine. Here quarantine is ended at *t =* 50 days, and then reinstated at *t =* 80 days. Quarantine then ends again at *t* = 150 days. Variables *E_Q_* and *E* are not plotted. Parameter values are those of Table 1.

## Results

Figures 2 and 3 are example simulations, to illustrate general model behavior, but are not optimal solutions. We now consider model projections, within our optimal framework. Our results are plotted from *t* = 0 days, where the initial conditions used are the estimated populations as of March 28th^14^.

### Gradual Release

The number of people to be released from quarantine, *M*_1_, was divided into a mesh of 1000 equally-spaced points ranging from 0 to the total quarantined population, *N_Q_* (0). Each one of these trial values for *M*_1_ was simulated against a mesh of 1000 equally-spaced points ranging from 0 to 400 for an associated release time *T*_1_. Once an optimum solution was found, a second optimum release pair, *M*_2_ individuals released at time *T*_2_ was also found. Unless specified otherwise, the base values used are 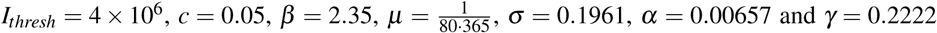 (these values are the same as presented in Table 1). The optimum solution was found for a range of values of *β*, *c* and *I_thresh_*, to observe how these uncertainties affect the optimum solution. These solutions are presented in Figure 4, where the top row (plots a to c) display the total quarantined population *(N_Q_)* under the optimum release strategy, and the second row displays the associated total infected population (*I* + *I_Q_*).

**Figure 4.**
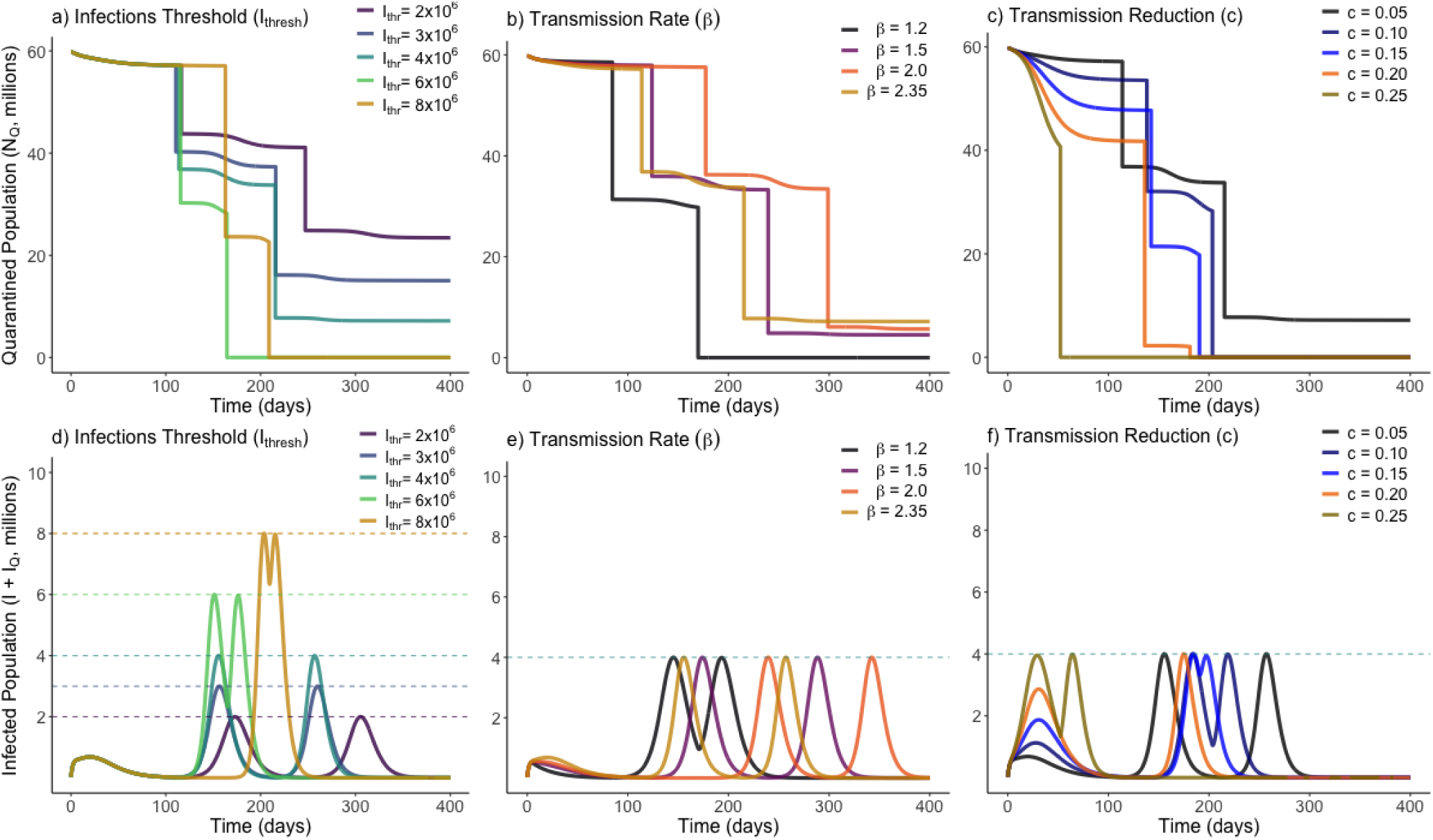
Optimum gradual release strategies for a range of different values of *I_thresh_* (infection threshold), *β* (transmission rate) and *c* (lockdown effectiveness), as marked. Plots a) to c) show the total quarantined population, displaying when releases from quarantine are made by the instantaneous decreases. Plots d) to f) depict the associated total infected population (*I* + *I_Q_)* associated with each optimum release strategy.

From Figure 4, we see that varying the infection threshold, *I_thresh_*, or the lockdown effectiveness, *c*, has the greatest impact on *M_i_* the number of people released, while the time of initial release *T_i_* remains mostly unchanged. This suggests that an increase of 1,000,000 to *I_thresh_* can allow approximately 4,000,000 more people to be released from quarantine. Changes to transmission (*β*) instead primarily adjusts the time at which releases are made, with the number of people released remaining relatively consistent. Figure 4b shows that for each trialled transmission value, approximately 50% of the quarantined population can be released once the current infected population reaches a sufficiently low level. In each case, there is approximately a two-week period between when the peak in infected individuals has ended and when individuals are released from quarantine. Figure 4c shows that, for less-effective lockdown measures, more individuals are able to be release from quarantine once the initial peak has ended. This seemingly counter-intuitive result is due to the reduced lockdown effectiveness meaning that a greater proportion of the quarantined population have been infected while in quarantine, and have since entered the recovered class. This means they can be re-added to the working pool without substantial risk of further infections. This result however clearly depends on the strength of any acquired immunity.

Additional to the graphical sensitivity analysis presented in Figure 4, a quantitative sensitivity analysis was also conducted on each model parameter. For each parameter, we calculated the total sensitivity index^25^, 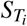, a variance-based sensitivity metric that shows how ‘important’ a model parameter is in affecting a certain model outcome. In this study, the model outcome considered is the objective function, *C*, for our optimum strategy. Defined formally, the total sensitivity index for parameter *i* is 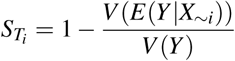, where *Y* is the model outcome monitored, and *X_i_* is the parameter considered. *X*_~_*_i_* here represents fixing all parameters except for parameter *i*. The total sensitivity index is equal to 1 minus the variance in the expected outcome when all parameters except the one in question are fixed, divided by the variance if no parameters are fixed.

In essence, for each model parameter a value between 0 and 1 is calculated that describes how sensitive the optimum release strategy is to that parameter, with a value nearer unity being more sensitive. Total sensitivity indexes were calculated for *β*, *σ*, *α*, *γ*, *c* and *I_thresh_*. In descending order, these were estimated through Monte Carlo sampling to be: *γ* : 0.4978, *β* : 0.3928, *I_thresh_* : 0.1994, *σ*: 0.0958, *c* : 0.0018, *α* : 0.0006. We therefore find that our optimum release strategy is not strongly dependent on the values of the disease-induced death rate, the lockdown effectiveness, or the incubation rate (*α*, *c*, or *σ*), and that in monitoring the effectiveness and outcome of a release strategy, the recovery rate and transmission rate of the disease should be most closely studied.

### On-Off Releases

To determine the optimal timings for an “on-off” lockdown release strategy, both the times at which quarantine was ended, 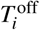, and the times at which it was reinstated, 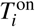, were iterated on a mesh of 500 evenly spaced points across a timespan of 0 to 400. Once one optimum release pair was found, the process was repeated up to two further times to identify subsequent optimum releases as necessary. Unless otherwise stated, the base values used were *I_thresh_* = 4 × 10^6^, *c* = 0.05, *β* = 1.5, 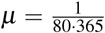, *σ* = 0.1961, *α* = 0.00657 and *γ* = 0.2222, which are again the same values listed in Table 1. A lower base value of *β* was used as it was considered unlikely that the population would be released from quarantine without certain social-distancing policies being implemented. The optimum solution was found for a range of different values of *β*, *c* and *I_thresh_*. These solutions are presented in Figure 5.

**Figure 5.**
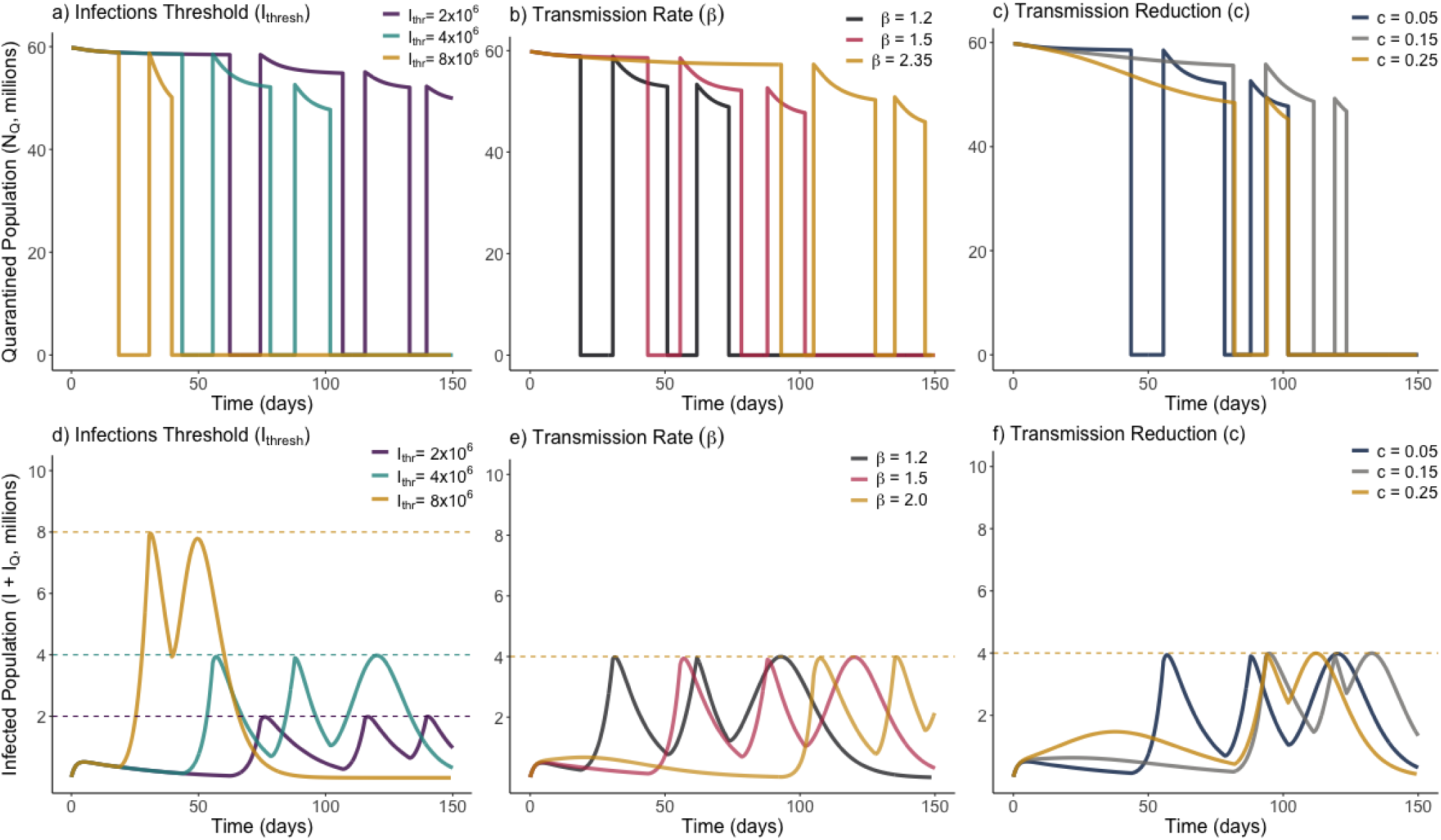
Optimum on-off release strategies for a range of different values of *I_thresh_* (infection threshold), *β* (transmission rate) and *c* (lockdown effectiveness), as marked. Plots a) to c) show the total quarantined population, displaying when releases and re-entry to quarantine are made. Plots d) to f) depict the associated total infected population (*I* + *I_Q_)* associated with each optimum release strategy.

From Figure 5 we see that, in all scenarios, it is never optimal to leave the entire population out of quarantine for long. Notably, we see in every instance that the optimum solution results in quarantine only being lifted for periods of 1–2 weeks. This presents a very narrow window of time to be able to monitor the rise in infections. There was no distinguishable change in dynamic behaviour between the different parameters sampled. We see in Figure 5b that increasing the transmission of the disease required the initial quarantine to be ended later, and for subsequent quarantines to be enforced for a longer duration. All optimum solutions do however result in rapidly increasing herd immunity by moving a large number of individuals to the recovered class. This difference can be seen by noting the difference in timescale on the horizontal axes of Figures 4 and 5.

Just as with the gradual release strategy, total sensitivity indices were calculated via the same method for our optimum on-off strategy. In descending order, and for the on-off strategy, these now become: *γ*: 0.6371, *I_thresh_*: 0.3775, *β*: 0.2973, *σ*: 0.2821, *c*: 0.0478, *α*: 0.0045. We see that the order of sensitivity is roughly the same as for the gradual release strategy, however all values (except for *β*) show increased sensitivity on the optimal result. The impact of the incubation period is the most substantially different, and this is due to the shorter durations of time people are out of quarantine in the optimum solution, meaning small changes to the incubation period may have large unexpected impacts on the surge in infected individuals.

## Discussion

Here we have investigated the optimal release of individuals from a state of lockdown. The primary conclusion of our work is that a gradual release strategy is far preferable to an on-off release strategy. We conclude this from the finding that a population-wide instantaneous release would cause the number of infected individuals to rise dramatically, in a short period of time. Any decision to begin easing lockdown measures will require constant monitoring and a high-level of population testing to track the likely rise towards a second-peak of infections. We show that employing a gradual release strategy, where groups of the population are slowly released from quarantine sequentially, will slow the arrival of any subsequent infection peaks compared to an on-off strategy, where lockdown is ended for all individuals imminently and reinstated when subsequent infections begin to increase. In all considered instances (i.e. parameter variations), it will not be possible to end lockdown for the entire population for any longer than two weeks, as the number of infected individuals is then expected to quickly overwhelm the health service following such a release. By ensuring that the increase in the number of infected individuals is as slow as possible, this will enable health officials to monitor more accurately the evolving situation, and provide more time to respond to unexpected increases in the number of infected individuals. We note that our approach does not consider the ethical responsibilities that will also impact any policy decision. If enough hospital provision was available, many more people can return to employment, but we recognise this will result in increased risk of further mortalities. As many governments state however, a functioning economy is more able to provide health provision to those with non-COVID19 life-threatening illness.

For a gradual release strategy, our simulations broadly suggest that a large section of the population should be released from lockdown initially, after the first peak of infections has fully passed. The rest of the population may then be released three to four months later following a likely second peak in infections. Again, in a general context, it is optimal to wait for one-to-two weeks after the end of an infection peak before releasing any of the population from lockdown. While it is desirable to return the population to work as early as possible, our optimal calculation states that this one-to-two week “wait” period is crucial in ensuring that the number of infected individuals is as low as possible when ending any lockdown measures, to reduce the growth of new cases. After this sufficient, cautious, wait period has ended, people should then be released from quarantine, with the knowledge that as many as 1 in 100 of them (under the worst-case scenario) may require critical care^10^ in the coming months. It is expected that a second peak in infections may be observed one to two months after this release date, and that the remaining population in quarantine should remain so until, once again, several weeks of low newly infected cases daily have been observed.

What we have not undertaken here, is to investigate or advocate any particular forms of changed behaviours that might be needed by those released, although understanding them can allow parameters (such as transmission rates) to be adjusted in our framework. Additional measures proposed include: reopening local connections before connecting cities further apart^26^, differential release times based on age^27–29^, on-going social distancing^9,30,31^, contact tracing using mobile applications^32^ and behaviour monitoring^33^, case-finding^34^ and cyclic schedules (e.g. short working weeks)^35,36^.

Placing our analysis in the context of other studies, Mulheirn et al. (2020)^34^ provide a particularly broad and qualitative assessment of ranges of possible exit strategies from lockdown. These include release times potentially dependent on age, sector, or geographical region, and the latter including metrics of local health capacity. Such measures can be in tandem with strong policies to shield the most vulnerable. The authors note that with varied approaches to lockdown release by differing countries, there is an opportunity to learn from this by intercomparison. Undoubtedly all countries leaving lockdown, however implemented, will heavily scrutinise data for any evidence of an emerging “second wave” of infections. Noted is the potential for raised levels of testing, in tandem with contact tracing for anyone found to be infected, to slow the spread of COVID-19 while at least a partial restarting of society occurs. For all of the options considered by Mulheirn et al. (2020)^34^, if the related parameters can be estimated with at least some certainty, then we believe our flexible model structure can adopt these. Hence our simulation framework provides a mechanism to place any suggested lockdown plans on a quantitative basis. Furthermore, where flexibility exists in release times, then for a given strategy, calculation of an optimal solution is possible.

The nearest analysis to ours found in the literature, based on both a SEIR framework and applied to COVID-19, is by German et al. (2020)^28^. Their version of the SEIR equations place more complexity into the infection component, differentiating between alternative levels of seriousness with which a person has the illness i.e. from asymptomatic through to requiring intensive care. They also allow for uncertainty as to whether people who recover are immune – an issue likely to be resolved once antibody tests become routinely available. Hence, people post-infection can, in the model, be returned to the susceptible pool. German et al. (2020)^28^ conclude that without retaining some constraints on the population after the termination of lockdown, then there would be an overwhelming increase in infections. Such constraints include social distancing, isolation of infectious people and contact tracing. They further stress the importance of a considerable increase in the testing of individuals to best inform any release decisions. Their conclusions align with many of our findings, however, rather than assessing constraints applied to the entire population as released simultaneously, our primary focus is to consider additional flexibility to constrain infection levels by a gradual release from lockdown.

Whilst we believe that our model framework does have predictive capability, we do raise a couple of caveats. We recommended exploring our findings within a variety of other model frameworks. Stochastic frameworks may be better suited to model the exact time periods when populations are first reintroduced, so as to better calculate the range of time frames until a second wave of infections in a probabilistic setting. Likewise compartmental infection models such as those presented by Giordano et al. (2020)^9^ will be able to provide more accurate estimations on any expected hospital intake.

In preparing to monitor the situation upon easing lockdown measures, our sensitivity analysis highlights that the recovery rate of the disease, *γ* above, is the most critical parameter in understanding the magnitude of any subsequent peaks in infection. Our calculations can be trusted further if that value is well-understood. For example, if new hospitalised patients of COVID-19 appeared to be remaining symptomatic and infectious for longer that previously estimated, it is plausible to assume within the general community that the disease is therefore being transmitted faster than previously expected. This knowledge could trigger preparations for a potential need to reinstate lockdown measures. Hence further research efforts into the infectious period should also therefore be prioritised. In a similar vain, the parameter to which results are second-most sensitive is transmission rate, *β*, and so also worthy of precise research.

A potential benefit of the on-off release strategy is that it greatly increases the number of people subsequently moved to the recovered class, rapidly bolstering the acquisition of herd-immunity. This in theory would enable the full re-opening of the economy at an earlier date, however it makes the critical assumption that recovered individuals would remain immune to the disease. The nature of immunity to SARS-CoV-2 is an open question and efforts are being made to understand its strength and longevity, but currently the WHO advises that there is no evidence yet to suggest that recovered COVID-19 patients have ongoing immunity to a second infection^37^. In light of this, the more cautious gradual release strategy remains even more preferable as the scientific community continues its efforts to develop a viable vaccine.

In conclusion, using an optimal control methodology, we have shown that a gradual staggered release of individuals out of lockdown is recommended to ensure that health systems are not overwhelmed by a surge in infected individuals. It has been well observed that older individuals are more likely to require critical care as a result of COVID-19^10^. Although our analysis does not as yet differentiate by age who should be in any partial lockdown releases, this does indicate that, potentially, the younger population could be the first to be released from lockdown. This would further ease any subsequent strain on the health system, and potentially further bolster a herd-immunity effect. Similarly, our analysis does not model the capability of businesses and individuals who have the infrastructure and availability to continue to work remotely.

The ongoing threat of COVID-19 will require continual monitoring and study in the coming months. It is important to ensure that infections are kept to a minimum, and that the government and relevant services are given enough time to prepare for increases in infections. The findings of this study stress that gradual and cautious action must be taken when easing lockdown measures, to save resources, and lives, while adding to the evidence base of possible routes out of lockdown.

## Data Availability

All associated code and data is made available at https://osf.io/hrt2k/.

## Acknowledgements

T.R. is supported through an EPSRC Systems Biology studentship award (EP/G03706X/1). T.R.B is supported by the BBSRC Industrial CASE studentship BB/R505067/1. C.H. gratefully acknowledges the Natural Environment Research Council award of National Capability Funds to the Centre for Ecology and Hydrology.

## Author contributions statement

T.R., C.H. and M.B.B. designed the model structure. T.R. performed the numerical simulations. D.V. and T.B. collated parameter values from the literature. T.B. produced the figures. All authors reviewed and contributed to writing the manuscript.

## Notes

### Competing Interest Statement

The authors have declared no competing interest.

